# Effectiveness of smartphone-based Community Case Management on urgent referral, re-consultation and hospitalization of children under-5 in Malawi: Results of a cluster-randomized, stepped-wedge trial

**DOI:** 10.1101/2020.09.03.20187328

**Authors:** Griphin Baxter Chirambo, Matthew Thompson, Victoria Hardy, Nicole Ide, Phillip H. Hwang, Kanika I. Dharmayat, Nikolaos Mastellos, Ciara Heavin, Yvonne O’Connor, Adamson S. Muula, Bo Andersson, John O’Donoghue, Sven Carlsson, Tammy Tran, Jenny Chen-ling Hsieh, Hsin-yi Lee, Annette Fitzpatrick, Joseph Tsung Shu Wu

## Abstract

**Background:** Integrated community case management (CCM) has led to reductions in child mortality in Malawi from illnesses such as malaria, pneumonia and diarrhoea. However, adherence to CCM guidelines is often poor, potentially leading to inappropriate clinical decisions and outcomes. We determined the impact of an electronic version of CCM (eCCM) application on referral, re-consultation and hospitalization rates of children presenting to village clinics in Malawi.

**Methods:** A stepped-wedge cluster-randomized trial compared paper-based CCM (control) with and without use of an eCCM app on smartphones from November 2016 to April 2017.A total of 102 village clinics from two districts in Northern Malawi were assigned to one of six clusters which were randomized to the sequencing of crossover from the control to the intervention phases, as well as the duration of exposure in each phase. Children ≥2 months to <5 years presenting with acute illness were enrolled consecutively by Health Surveillance Assistants (HSAs). The primary outcome of urgent referrals to higher-level facilities was evaluated using multi-level mixed effects models. A logistic regression model with random effect of cluster and fixed effect for each step was fitted. Adjustment for potential confounders included baseline factors, such as patient’s age, sex, and geographical location of village clinics. Calendar time was adjusted for in the analysis.

**Results:** A total of 6965 children were recruited, 3421 in the control and 3544 in the intervention phase. After adjusting for calendar time, children in the intervention phase were more likely to be urgently referred to a higher-level health facility compared with children in the control phase (OR 2.02, 95% CI 1.27-3.23; p<0.01). Overall, children in the intervention arm had lower odds of attending a repeat HSA consultation (OR 0.45, 95% CI 0.34-0.59; p<0.01) or hospital admission (OR 0.75, 95% CI 0.62-0.90; p<0.01), but after adjusting for time these differences were not significant (p>0.05).

**Conclusions:** Addition of eCCM decision support led to a greater proportion of children being referred to higher-level facilities, with no apparent increase in hospital admissions or repeat consultations in village clinics. Our findings provide support for the implementation of eCCM tools in Malawi and other Low and Middle Income Countries (LMIC), with a need for ongoing assessment of effectiveness and integration with national digital health strategies.

**Trial registration:** http://ClinicalTrials.gov; NCT02763345. Registered 3 May 2016

## Background

Community Case Management (CCM) is a paper-based clinical decision aid that enables frontline community health workers (CHW) to recognize and manage the major causes of mortality among children under-five in low and middle income countries (LMICs), specifically malaria, pneumonia, diarrhea, and malnutrition(1). CHWs, known as Health Surveillance Assistants (HSAs) in Malawi, deliver CCM from village clinics situated in hard-to-reach locations. In a country where 35% of the population reside in rural locations more than 8km from facility-based health providers, CCM has helped provide equitable access to healthcare for many communities that were previously underserved(2). CCM is widely implemented in LMICs and has demonstrated some success in improving the management of children with acute illness and reducing mortality. For example, CCM of pneumonia reduced pneumonia-associated mortality by 70% in developing countries(3)(4). Since adoption of CCM in Malawi in 2008, this strategy has contributed to significant reductions in under-five mortality from 97.8 per 1,000 live births to 52.7 in 2017(5).

Implementation of CCM is based on the diagnostic and prognostic value of certain signs and symptoms; children with uncomplicated illness are managed in the community with symptomatic treatment, while those with more worrisome ‘danger’ signs are urgently referred to a health facility that can provide a higher-level of care. Accurate and timely urgent referral is an integral component of CCM and is a prerequisite for improving health outcomes among acutely unwell children. Aside from preventable mortality, failure to refer at the appropriate time can over-burden constrained resources at higher-level facilities needed for treatment of severely unwell children, lead to unnecessary hospital admissions, or involve caregivers repeating inconvenient journeys to village clinics because their child’s condition has not resolved or is deteriorating.

When implemented appropriately, CCM is an effective childhood survival strategy, but there is considerable evidence of suboptimal management decisions from field studies, including poor HSA adherence to the required steps of CCM (6). As CCM is delivered using a paper-based tool, it is easy for relevant fields to be over-looked or for clinically invalid data to be recorded, contributing to inappropriate management decisions. Poor adherence to CCM is further compounded by structural and sociocultural factors such as transportation costs or distance to higher-level facilities that further impede potential impact of CCM.

The proliferation of mobile phones and cellular networks in LMICs has prompted development of several mHealth technologies as alternative platforms for delivering childhood intervention strategies (7). Decision support systems within these technologies can facilitate clinical decision making, and offer potential advantages over paper-based tools(8)(9). However, adopting mHealth tools as an adjunct (or replacement) for current paper-based assessment and reporting tools requires considerable initial and sustained investment. Therefore, demonstrating evidence for the impact of mHealth on health care outcomes is crucial. We determined the impact of an electronic version of CCM delivered as a smartphone app, compared to standard delivery of CCM on outcomes of referral rates and hospital admissions of children under 5 years presenting to village clinics with acute illness in Malawi.

## Methods

### Study design

The full study protocol has been published previously (10). In summary we used a stepped-wedge cluster-randomized design trial to compare paper-based CCM (paper CCM) with a mobile phone application version of CCM that was developed by the Supporting LIFE research team (SL eCCM App) (11). Village clinics were grouped into six clusters based on geographic proximity, and clusters were randomized to determine the sequence of crossover from the control (using paper CCM alone) to the intervention (using paper CCM as well as SL eCCM), and the duration of exposure (between 2-7 weeks) in each phase. Children aged ≥2 months to <5 years were triaged using paper CCM or paper CCM + SL eCCM depending on when they presented to village clinics.

This study was conducted in 102 village clinics across Nkhata Bay and Rumphi districts in Northern Malawi where Chichewa, Chitumbuka and Tonga are the principal languages spoken. Cassava farming is the main occupation of the 215,429 inhabitants of Nkhata Bay District, situated on Lake Malawi. Rumphi district, which extends west from Lake Malawi to the Zambian border, is principally a tobacco farming community with 166,460 inhabitants. According to National Statistic Office of Malawi, 23.4% and 17.4% of the adult population (defined as aged 15 years and above) are illiterate in Nkhata Bay and Rumphi, respectively. Village clinics are typically basic community structures equipped only with rudimentary diagnostic aids (e.g. stopwatch) and medical supplies (e.g. oral rehydration therapy, artemisin-based combination therapy). Each clinic is operated by a single HSA who is a government-employed CHW responsible for assessing and managing acutely unwell children using CCM. HSAs are people who possess a Malawi Schools Certificate of Education (MSCE) or Junior Certificate of Education (JCE). They are the frontline health workers who work in the village clinic(12). A village clinic serves a catchment area of approximately 1,000 people(13)(14). Both districts are predominantly rural with 30-35% of the population residing more than 8km from a health facility with qualified clinical staff(15).

### Participants

A total of 102 HSAs were recruited to participate (77 male, 24 female, 1 not disclosed), with age ranging from 27 to 59 years, who had been working as HSAs for between 1 and 27 years (mean 10.1 years). Prior to the trial, 101 HSAs already had phones, of which 37 were smartphones.

HSAs consecutively enrolled eligible children aged ≥2 months to <5 years from November 2016 to February 2017, who presented with acute illness, but were not unconscious, convulsing, or previously enrolled in the study. Due to high rates of illiteracy in Nkhata Bay and Rumphi Districts, caregivers were required to verbally consent in their preferred language for their child to be enrolled to avoid limiting participation by literacy levels.

### Randomization and blinding

Village clinics were grouped into six clusters based on their geographic proximity. Clusters were then randomized using an online random number generator to determine the sequencing of crossover of clusters from the control phase (paper CCM) to the intervention phase (paper CCM + SL eCCM), as well as the duration of exposure (between 2-7 weeks) in each phase. Neither participants nor researchers were blinded to allocation. Baseline data was collected during the index visit and recorded in the village clinic register (VCR) in the control arm and additionally in the SL eCCM app during the intervention phase.

### Health Surveillance Assistants (HSAs) Training

Before commencing recruitment, we engaged with national and district level health authorities to explain to them the objectives of the clinical trial and the procedures to be done. HSAs recruited by the study team attended a 1-day training workshop to learn study procedures for the control and intervention phases of the trial, as well as how to operate the SL eCCM App and the smartphone (details of the hardware and development and functionality of the App are described in full elsewhere(16)). Prior to crossover from the control to the intervention phase, HSAs attended a further 2-day training workshop to re-familiarize them with study procedures and the technology specific to the intervention phase. Training workshops were conducted in English with a member of the research team fluent in the regional dialects of Chichewa, Tonga and Chitumbuka present to facilitate communication. HSAs were permitted to use their clinical discretion if they disagreed with care recommendations.

### Procedures

During the control phase, HSAs used paper-based CCM to assess and manage children for 2 to 7 weeks (depending on the assigned order of clusters after randomization). Details of clinical presentations were recorded by hand in the VCR, as per standard practice. During the intervention phase, HSAs recorded clinical presentations synchronously in both the VCR and the SL eCCM app. Form validation ensured that HSAs were required to complete every field on the eCCM App(17). A complete inventory of the data collected by HSAs during each phase of the trial is reported in full elsewhere(10). Outcome data was collected retrospectively using local nursing students/graduates who travelled to village clinics and higher-level health facilities within an assigned catchment area on a weekly basis to manually abstract data from patient records. Enrolled children were re-identified at clinical sites by cross-referencing each child’s full name, date of enrollment, date of birth, and sex along with caregiver name and contact details pre-populated on clinical research forms. Given that caregivers can re-present to any village clinic and higher-level health facility, child identifiers and subsequent attendances at health facilities were verified by caregivers via cell-phone beforehand, to improve efficiency of data collection. Clinical data was recorded in the VCR during both control and intervention phases of the study in order to fulfill local reporting requirements for village clinics and the district health authorities.

### Outcomes

The primary outcomes were urgent referrals to higher-level health facilities, re-consultations to village clinics, and hospitalization within 7-days of study enrolment. Outcomes were determined by case note review of VCR and health facility records undertaken 2-weeks after study enrolment. Re-consultations were defined as re-attendances to any village clinic for a health concern related to the reason for presentation recorded in the VCR at baseline, and hospital admissions related to an inpatient stay in a secondary or tertiary care facility for ≥1 day due to deterioration of illness recorded in the VCR.

### Statistical analysis

Descriptive analyses examined individual-level baseline characteristics of children between the control and intervention periods across all clusters. Participant characteristics included the child’s age, sex, district in which the child was recruited, and the treatment decision by the HSA at the index visit. For continuous measures, we reported means and standard deviations. For categorical measures, we reported counts and proportions. *t* tests and χ^2^ tests were used to determine bivariate differences for continuous and categorical measures, respectively.

Following an intention-to-treat principle, we evaluated overall differences in the proportion of children who were urgently referred to a higher-level health facility by HSAs between the control and intervention periods. For the primary analysis, urgent referrals to higher-level facilities were evaluated at the patient-level using multi-level mixed effects models(18). Since all outcomes were binary, a logistic regression model with random effect of cluster and fixed effect for each step was fitted(19)(20). Potential confounders included baseline factors of, age, sex, and geographical location of village clinics (i.e. urban/rural). Calendar time was also adjusted for in the analysis and the effect of time was modeled as a categorical variable(19). Odds ratios (ORs) with 95% confidence intervals (CIs) were estimated and reported. Potential effect modification by age and sex were assessed by stratification as well as inclusion of interactions terms with the intervention period in the model. Other main outcomes, of re-consultations at village clinics and hospitalizations, were also analyzed using mixed models with the same potential confounders and effect modifiers. For secondary analyses, we explored for heterogeneity in treatment effects between clusters, using within-cluster comparisons between the intervention and control periods. A p-value of <0.05 was considered statistically significant and all tests were two-sided. All analyses were conducted using Stata version 14 (StataCorp, College Station, Texas). Patient flow, including the number and reasons for exclusion and withdrawal were reported according to CONSORT recommendations.

## RESULTS

### Participants recruited and baseline characteristics

A total of 6965 children were recruited, of whom 3421 were included in the paper CCM (control) phase, and 3544 in the eCCM (intervention) phase (Table 1). A larger number of children were recruited from the Nkhata Bay district than Rumphi in both trial phases reflecting the higher population in the former, as noted above. Children recruited in the control phase were significantly younger (mean age 24.6 months), than those in the intervention phase (26.9 months). The majority of children in both the control (3217 or 94.0%), and in the intervention phases (3335 or 94.1%) were treated at home after their assessment by the HSA (Figure 1).

### Effect of intervention on urgent referral

A total of 179 children (5.2%) in the control phase were urgently referred to a higher-level health care facility, compared to 198 (5.6%) in the intervention phase (OR 1.04, 95% CI 0.83-1.29; p<0.75) (Table 2). After adjusting for the effects of time (Model 1), the OR was 2.02 (95% CI 1.27 to 3.23; p<0.01) in favor of the intervention. Further adjustment for age, sex, and district (Models 3 and 4) provided similar strengths of association. We found no significant effect modification with the models tested. Examination of cluster-specific treatment effects, adjusted for calendar time, across the 6 clusters showed that the direction of effect was identical to the overall effect (i.e. favoring intervention) in 4 of the 6 clusters, but was statistically significant in only one of these (cluster 2, p<0.01) most likely affected by sample size. In 2 of the 6 clusters (clusters 3 and 4) the direction of effect favored the control arm, but was statistically significant in only one of these clusters (cluster 4, p<0.01).

### Effect of intervention on repeat consultation

Repeat consultations with an HSA within 7 days occurred in 180 (5.3%) children in the control phase, compared to 86 (2.4%) in the intervention phase (OR 0.45, 95% CI 0.34-0.59; p<0.01) (Table 2). After adjusting for the effects of time (Model 1), the OR was 0.57 (95% CI, 0.32-1.04) in favor of intervention arm, but not statistically significant (p = 0.07). Further adjustment for age, sex, and district (Models 3, 4) provided similar ORs. We found no significant effect modification with these models. Examination of cluster-specific treatment effects showed that the direction of effect for the outcome of repeat consultation favored the control arm in four clusters (cluster 1, 3, 4 and 5) and was statistically significant in only one of these (cluster 4, p = 0.04), while in clusters 2 and 6 it favored the intervention, but was not statistically significant (p>0.05).

A total of 219 clinical features were recorded on the children attending repeat consultations, although not all children had clinical symptoms recorded and more than one clinical feature could be recorded on one child. The most common symptoms reported were fever for <7 days, cough for 21 or more days, diarrhea <14 days without blood, and fast breathing (Table 3).

### Effect of intervention on hospital admission

A total of 565 children were admitted to hospital, 320 (9.4%) in the control arm, and 245 (6.9%) in the intervention arm (OR 0.75, 95% CI 0.62-0.90; p<0.01) (Table 2). Of these, 290 (51.3%) were urgently referred by their HSA, and 270 (47.8%) taken to hospital by their parents (information on origination was not available on 5 children). After adjusting for the effects of time (Model 2), the OR was 1.23 (95% CI 0.83-1.81) in favor of the control arm but was not statistically significant (p = 0.30). Further adjustment for age, sex, and district (Models 3, 4) provided similar ORs. We found no significant effect modification with these models. Examination of cluster-specific treatment effects showed that the direction of effect on hospital admission favored the control arm in four clusters (cluster 1, 2, 5 and 6) and was statistically significant in only one of these (cluster 2, p<0.01), and the intervention arm in two clusters (3 and 4), but was not statistically significant (p>0.05).

## DISCUSSION

### Main findings

We found that the addition of eCCM to the usual practice of assessing and treating children by HSAs using the paper based assessment tool led to a significant increase in the proportion of children referred urgently to higher-level health care facilities. This direction of effect of the intervention was found in most but not all of the 6 clusters of clinics. The intervention was also associated with smaller proportions of children who attended for a repeat consultation at the village clinic, or who needed to be admitted to hospital, but these were not statistically significant after full adjustment in models.

We speculate that the modest effect of eCCM on increasing the proportion of children who were urgently referred may have been due to greater adherence to the CCM decision support algorithm(21,22), as the smartphone app encouraged adherence to CCM(22), and/or that the smartphone app reduced errors by providing more accurate assessment than paper CCM alone. The effects could, however, merely represent the impact of the requirement in our trial for HSAs to double enter data (i.e. on paper format and then using the app) with replication of assessment providing more opportunity to conduct the required assessment. Children in the intervention group were less likely to return for repeat consultation, which could be due to their assessment being more thorough and/or correct at their initial visit to the village clinic, thus reducing the need to return to the clinic for the same illness.

Our trial demonstrates that the vast majority of children under 5 years of age presenting with acute illness are managed in the community by HSAs, with only approximately one in twenty being urgently referred to a higher-level health care facility. Some caregivers appear to bypass the village clinic and attend the hospital directly without first being assessed at the village clinic. This implies dissatisfaction with the services provided by lower level health care facilities such as village clinics (13). One reason that caregivers may bypass village clinics and go directly to higher level facilities may be because caregivers perceive that their child needs urgent attention and may feel that going to the village clinic wastes time(23). Indeed, previous studies have found that some community members do not have confidence in the care they get from the first level health care providers such as community health workers(24,25).

A major strength of our study is the use of a rigorous stepped-wedge design, which aimed to minimize potential biases. Previous research has established the considerable support by HSAs and the local community for the planned intervention(12,14,26). We could not conduct an individual randomized study at the child level, nor at the HSA level, following guidance from local investigators. A stepped-wedge design has significant advantages over a simple before/after design as it attempts to minimize potential temporal biases and provides a more powerful study design using a control group. We selected referrals as the main trial outcome as this is a major driver of use of health care utilization and there is currently evidence that under-referral contributes to child morbidity(27)(28). We believe that this study provides a high degree of generalizability to similar settings in Sub-Saharan Africa where CCM is used.

This study does have some limitations. One involved the requirement of our study design for HSAs to continue to use the paper tool during the intervention phase, which means we could not assess the independent impact of eCCM, but rather its added value. This occurred because the SL eCCM App was not yet endorsed by the Malawi Ministry of Health so it could not replace the paper tool. In addition, assessing outcomes was challenging due to lack of unique patient identifiers and incomplete records at health care facilities. The study may have been under-powered to detect effect on repeat consultations and hospital admission, and we noted heterogeneity between clusters in direction and significance of effects.

Our results add to previous qualitative studies which have shown that HSAs feel empowered using apps such as the one evaluated here (29), and that these tools might support greater adherence to CCM(21). These results support wider implementation and potentially addition of further functionalities, to support HSAs using smartphones to facilitate other tasks, such as reporting requirements or drug stocking. For policy makers, our study provides robust evidence on the effectiveness of this eCCM tool that few other studies have provided to date, and could therefore add evidence to support national digital health strategies in developing an integrated community health information system. However, we acknowledge that further data is needed to determine costs (and cost effectiveness) of initial set up (training, smartphones, servers), and ongoing support (technical support, hardware replacement, software updates). For researchers in this area, evidence for the independent effects of eCCM on outcomes is needed. These results call for additional studies on eCMS applications and similar mHealth tools to determine impact on other clinical outcomes such as duration of illness and resource utilization. In addition, there is a need to ensure data and workflow integration of eCCM as part of community health services delivery, management and existing digital tools used at other levels of health system, such as the District Health Information System (DHIS), electronic medical records systems and real time surveillance at the community level.

## Conclusions

The vast majority of children assessed by HSAs in these two large rural areas of Malawi are managed by HSAs in their communities. Adding an electronic version of CCM that involved an app on smartphones, to HSAs’ usual paper-based CCM tool, led to a significant increase in the proportion of children referred from village clinics to higher level health care facilities. While effects on decreased hospital admission, and decreased repeat consultation were suggestive but inconclusive, they support the hypothesis that the eCCM tool improved decision making at the HSA level. Combined with existing qualitative literature showing high levels of acceptability of mHealth versions of CCM by community health workers, our findings support further efforts to deploy smartphone based tools as part of integrated digital health strategies in Malawi and similar countries in sub-Saharan Africa, with ongoing evaluation of effectiveness, cost-effectiveness, and acceptability by health care workers and community members.

## Data Availability

Data will be made available to anyone upon request to the College of Medicine Research Ethics Committee (COMREC).

## List of abbreviations

CCM: Community Case Management
CHW: Community Health Worker
CRF: Case Report Form
DHIS: District Health Information System
HSA: Health Surveillance Assistants
LMIC: Low and Middle Income Countries
OR: Odds Ratio
SL eCCM: Supporting LIFE electronic Community Case Management
VCR: Village Clinic Register

## Declarations

Ethics approval and consent to participate: The study received ethical approval from the University of Washington Human Subjects Division in Seattle USA (#51750), Imperial College of London Research Ethics Committee (16IC3396) in London UK and the College of Medicine Research and Ethics Committee (COMREC) in Blantyre Malawi (P.07/16/1984). National and community-level permissions were also obtained prior to the conduct of this study.

## Consent for publication: N/A

Availability of data and materials: The datasets used and/or analyzed during the current study are available from the corresponding author on reasonable request.

## Data Sharing

The results of this study will be disseminated through presentations in research conferences and copies of this study will be put in the Libraries of Mzuzu University, College of Medicine, University of Washington and University College Cork.

## Authors’ contributions

GBC helped in development of the questionnaire and interview guide, data collection, and writing the manuscript. NM and TT contributed to trial design and management and commented on this manuscript. KD contributed to the conduct of the study and commented on this manuscript. JOD conceived the study, developed the research question, participated in trial management and commented on this manuscript. PH developed the statistical analysis plan, and analysed and interpreted the data. NI managed field data collection and contributed in writing the manuscript. JTSW assisted in the study implementation, communication with national and district health authorities, and commented on this manuscript. All authors contributed to interpretation of findings, drafting of the manuscript and approved the final version.

## Acknowledgements

The authors would like to thank University of Malawi (College of Medicine), Mzuzu University, University of Washington and Imperial College London for giving us permission to conduct this study.

